# Standardisation of terminology, calculation and reporting for assigning exposure duration to drug utilisation records from healthcare data sources: the CreateDoT framework

**DOI:** 10.64898/2026.02.18.26346576

**Authors:** Judit Riera-Arnau, Olga Paoletti, Rosa Gini, Nicholas H Thurin, Patrick C Souverein, Shahab Abtahi, Carlos Durán, Romin Pajouheshnia, Giuseppe Roberto

## Abstract

**Background:** In pharmacoepidemiological studies, days of treatment (DoT) duration associated with individual electronic drug utilization records (DUR) are usually missing. Researcher-defined duration (RDD) calculation approaches, as opposed to data-driven approaches, can be used to estimate DoT based on the specific choices and assumptions made by investigators. These are usually underreported or even undocumented. We aimed to develop a framework for the standardization of terminology, formulas, implementation, and reporting of possible RDD approaches.

**Methods:** A systematic classification of RDD calculation approaches was developed via expert consensus. Universal concepts used to operationalise RDDs were identified and described using standard terminologies. An open-source R function, CreateDoT, was created to implement the formulas universal concepts as input parameter. A step-by-step workflow was developed to facilitate implementation and reporting.

**Results:** RDD approaches were classified in two main classes: I) daily dose (DD)-based calculation approaches (n=3 formulas), and II) fixed-duration approaches (n=2). Seven universal concepts were identified to describe the five corresponding generalized formulas for DoT calculation. Input parameters of the CreateDoT function can be retrieved from source data through its mapping to universal concepts, or inputted by the investigator based on the chosen calculation approach. The input file structure itself represents a standard reporting template for documenting investigators’ assumptions and methodological choices adopted for DoT calculation.

**Conclusions:** The CreateDoT framework can facilitate the documentation and reporting of RDD approaches for DoT calculation, increasing transparency and reproducibility of pharmacoepidemiological studies regardless of the data model used, and facilitates sensitivity analyses to evaluate the impact of alternative assumptions in DoT calculation.

## 1. BACKGROUND

In longitudinal pharmacoepidemiology studies based on secondary use of real-world healthcare data sources, the exposure to medication is typically assessed through drug utilisation records (DUR) based on the prescribing, dispensing or administration of medicinal products [1–4]. For each DUR of interest, a start date and an end date of treatment are identified. Treatment start date is usually assumed to correspond to the recorded date of drug prescription, dispensing or administration, while the end date is assigned based on the days of treatment associated with the DUR. Start and end dates of patient-level longitudinal sequences of DURs can then be used to build episodes of continuous treatment according to the study-specific exposure definition [5]. However, the number of days of treatment (DoT) corresponding to the prescribed, dispensed or administered medication is a variable that is often not collected or is partially missing [6] in real-world healthcare data sources and, therefore, needs to be estimated and imputed for the DUR(s) of interest.

Meiadi and colleagues classified methodological approaches used to impute DoT of a DUR in two main categories: 1) data-driven and 2) researcher defined duration (RDD) approaches [7]. In general, data-driven approaches estimate the number of days of exposure based on the observed distance between DURs of interest. These methods have the advantage of automating the process of estimating treatment duration, although they only provide reliable results when applied to treatments with a regular dosing schedule (e.g., long-term use) [8–10]. RDD approaches, instead, are based on *ad hoc* assumptions and/or computational formulas that are defined by the investigator considering the information available in the data together with other context information, such as prescribing recommendations, dosing regimens and expected utilisation patterns of the drug(s) of interest in the study population [11–14].

Indeed, different assumptions and methodological choices for estimating DoT using real world data can significantly affect study results [15–17]. As for RDD approaches, a recent study showed that, even keeping the calculation formula constant, different assumptions on daily dose and prescribed quantity resulted in a substantial difference in terms of estimated person years exposed to the study drug, number of exposed events and risk ratios [15]. RDD approaches, with relevant calculation formulas and assumptions about input parameters, need to be tailored to the specific data and study with the aim of obtaining the most accurate representation of the actual pattern of drug exposure in the study population [18]. In this context, the transparent reporting of methodological choices for DoT estimation based on RDD approaches becomes fundamental for facilitating reproducibility and interpretation of study findings [7,9,10,19]. Nevertheless, to the best of our knowledge, there is no systematic classification of RDD approaches available in scientific literature. This hampers both the use of a common reporting terminology for the reporting of methodological choices for DoT calculation and the development of statistical programs to standardise their implementation. Therefore, the aim of this study was to systematically classify RDD approaches to estimate DoT in real-world healthcare data sources and consequently develop an open-source R function to standardise their implementation.

## 2. METHODS

### 2.1 Systematic classification of Researcher Defined Duration (RDD) approaches

A comprehensive list of RDD calculation approaches for the calculation of DoT corresponding to a single DUR were identified by the study team (GR, NT, JRA, RP, OP, PS, RG, SA, CD) based on extensive experience in pharmacy, clinical pharmacology, pharmacoepidemiology, healthcare databases, data engineering, drug utilisation research and multi-database studies. RDD calculation approaches were then generalised and grouped according to a systematic classification. Universal concepts needed to describe and operationalise RDD calculation approaches and corresponding computational formulas were identified and defined using, whenever possible, the standard terms of the European Directorate for the Quality of Medicine (EDQM) and International Organisation for Standardisation (ISO) standards of the European Normalisation Committee for the Identification of Medicinal Products (IDMP) [20,21].

### 2.2 Creation of an R function for standardising the calculation of exposure duration

An open source R function named “Create Days Of Treatment” (*CreateDoT*) was developed to standardise the computation of DoT associated with DURs according to various RDD calculation approaches and corresponding computational formulas. The function was designed to also provide as a secondary output the corresponding average amount of active substance that would have been taken daily (i.e., daily dose measured as amount of active substance), according to either the calculated DoT or the recorded DoT whenever available in the data. Input parameters were defined according to the concepts used to describe and operationalise the different RDD calculation approaches with the aim of designing a function that can work with any data model.

As an example of CreateDoT application on a specific data model, concepts describing the input parameters of the function were projected to the ConcepTION Common Data Model (CDM) [22,23]. With this specific purpose, variables from the MEDICINES and PRODUCTS tables of the ConcepTION CDM were used [23] (see **Supplementary Table 1**). Additionally, example input and output template shell tables for CreateDoT implementation and reporting, based on the ConcePTION CDM, are provided in Additionally, example tnput and output template shell tables for CreateDoT implementation and reporting based on the ConcePTION CDM are presented in Supplementary Table 2.

The *CreateDoT* function was developed using R version 3.4.1 [24] and belongs to the CreateDoT R-package. It is stored in the following open access GitHub repository: https://github.com/IMI-ConcePTION/CreateDoT/wiki/ [25].

### 2.3 Examples of the application of the *CreateDoT* function

Dummy examples were created to illustrate the application of different RDD calculation approaches of the *CreateDoT* function in the context of three different case scenarios including the use of I) acitretin for psoriasis, II) apixaban for stroke and III) valproate for epilepsy, exemplifying three different levels of granularity of the assumptions, four different calculation approaches, and some variations on input data availability.

## 3. RESULTS

### 3.1 Systematic classification of Researcher-Defined Duration (RDD) calculation approaches

RDD calculation approaches were classified in two main classes corresponding to: 1) three daily dose (DD)-based calculation approaches and 2) two fixed duration-based calculation approaches. The distinction between the two classes is based on the type of information that the researcher is expected to input and make a decision upon.

A total of seven concepts corresponding to the parameters necessary to calculate DoT with the proposed RDD calculation approaches were identified and defined as reported in **Table 1**.

**Table 1.** Definitions of concepts used to describe RDD calculation approaches.

| Concepts | Definition* | Examples |
| --- | --- | --- |
| **Drug utilisation record (DUR)** | A record that stores information on a prescription, dispensing, or administration event of a specific drug. The record might contain one or more packages of the same medicinal products. | A patient receives on day X a prescription of two packages of paracetamol 1g of 32 tablets along with a bottle of acetylcysteine syrup 2% in 100ml. This event would trigger the creation of two distinct DURs:<br>I) DUR(1): two packages of paracetamol 1g, 32 tablets<br>II) DUR(2): one bottle of 100ml acetylcysteine syrup, 2%<br>Depending on the intrinsic characteristics of each DUR, different types of information might be available/recorded about the prescribed/dispensed/administered medicinal product. |
| **Medicinal product package<sup>&</sup>** | A medicinal product authorised for marketing, containing substances to treat, prevent, or diagnose diseases. Identified at the package level by a national/international medicine identifier code. | 1. 'TENORMIN® (atenolol) 50 tablets 50 MG'<br>2. 'LUMIGAN® (bimatoprost) eye drops 30 single-dose containers 0,4ML 0,3MG/ML'<br>3. 'DICLOREUM® (diclofenac) 30g cream tube' |
| **Unit of presentation** | Qualitative terms identifying the countable entity in which the strength(s) of the medicinal product is presented. | 1. 'TENORMIN® (atenolol) 50 tablets 50 MG', unit of presentation: tablet.<br>2. 'LUMIGAN® (bimatoprost) eye drops 30 single-dose containers 0,4ML 0,3MG/ML', unit of presentation: single-dose container<br>3. 'DICLOREUM® (diclofenac) 30g cream tube', unit of presentation: tube |
| **Active substance** | Any substance or mixture of substances contained in a medicinal product that exerts the intended pharmacological, immunological or metabolic action with a view to restoring, correcting or modifying physiological functions or to make a medical diagnosis. | 1. 'TENORMIN® (atenolol) 50 tablets 50 MG', active substance: ramipril<br>2. 'LUMIGAN® (bimatoprost) eye drops 30 single-dose containers 0,4ML 0,3MG/ML', active substance: bimatoprost<br>3. 'DICLOREUM® (diclofenac) 30g cream tube', active substance: diclofenac |
| **Pharmaceutical product** | Qualitative and quantitative composition of a medicinal product in the pharmaceutical dose form approved for administration in line with the regulated product information. | 1. 'TENORMIN® (atenolol) 50 tablets 50 MG', pharmaceutical product: (atenolol) tablets 50 MG'<br>2. 'LUMIGAN® (bimatoprost) eye drops 30 single-dose containers 0,4ML 0,3MG/ML', pharmaceutical product: (bimatoprost) eye drops 0,3MG/ML'<br>3. 'DICLOREUM® (diclofenac) 30g cream tube, 1%', pharmaceutical product: (diclofenac) cream, 1% |
| **Daily Dose (DD)** | It is the prescribed or assumed dose of a medicinal product taken daily. It can be measured either as the number of units of presentation, amount of active substance or amount of pharmaceutical product. | 1. number of units of presentation: e.g., DD=1 tablet of atorvastatin.<br>2. amount of active substance: e.g., DD=150 mg of metoprolol.<br>3. amount of pharmaceutical product: e.g., DD=10 ml of syrup. |
| **Fixed duration** | It is the prescribed or assumed number of days of treatment associated to either the DUR or the medicinal product package of interest. | It can be assumed by the investigator based on the expected prescribing behaviours and/or dispensing or even reimbursement recommendations/restrictions, e.g.:<br>1. Dispensing of isotretinoin cannot exceed 30 days of duration in women of childbearing age.<br>2. Prescription/dispensing of an antibiotic treatment for a defined number of days, irrespective of the strength.<br>It may also be available in the data as prescribed treatment duration or days supplied. |

DD: daily dose; DUR: drug utilisation record
\*The definitions listed here were created to align with the document *Standard Terms of the European Directorate for the Quality of Medicines (EDQM) [20]* and the corresponding items from the *ISO standards for the Identification of Medicinal Products (IDMP) [21]*.
&Brand names have been taken from the *Spanish Agency of Medicines and Sanitary Products (AEMPS) [26]*.

In **Table 2**, formulas corresponding to each RDD calculation approach are described using the identified concepts.

**Table 2.** Research Defined Duration calculation approaches and corresponding formulas for Days of Treatment (DoT) calculation.

| DD-based calculation approaches |  |
| --- | --- |
| Unit of presentation per day | DoT = total number of units of presentation in the DUR / DD |
| Active substance amount per day | DoT = total active substance amount in the DUR / DD |
| Amount of pharmaceutical product per day | DoT = total amount of pharmaceutical product in the DUR / DD |
| Fixed duration-based calculation approaches |  |
| Fixed record duration | DoT = fixed duration of the DUR |
| Fixed medicinal product package duration | DoT = number of medicinal products package units in the DUR * fixed duration of the medicinal product package |

DD: daily dose; DoT: days of treatment; DUR: drug utilisation record

#### 3.1.1 DD-based calculation approaches

The implementation of DD-based calculation approaches require the investigator to input a DD. The latter might be already available in the data (i.e., prescribed daily dose [PDD]), or imputed based on the investigators’ assumption of the expected use of the drug, or even based on already available reference standards, such as the Defined Daily Dose (DDD) set by the World Health Organisation (WHO) [14,27]. The DD-based class can be further distinguished based on the unit of measurement of the DD chosen by the investigator, that is:

1. units of presentation per day (e.g., DD=1 tablet of atorvastatin), for which DoT are calculated based on the total number of units of presentation in the DUR, divided by the number of units of presentation to be taken daily according to the prescribed or assumed DD (e.g., 2 tablets per day).
2. amount of active substance per day (e.g., DD=150 mg of metoprolol), for which the total amount of active substance corresponding to the content of the DUR is divided by the amount of active substance prescribed or assumed as DD. Notably, the total amount of active substance contained in the DUR of interest might be not readily available in the data and needs to be calculated. See section “*Computing the total active substance amount per medicinal product*” for more details.
3. amount of pharmaceutical product per day (e.g., DD=10 ml of cough syrup), for which DoT of a DUR is calculated by dividing the total amount of pharmaceutical product by the amount of pharmaceutical product to be taken daily according to the prescribed/dispensed or assumed DD

#### 3.1.2 Fixed duration-based calculation approaches

The implementation of the second class of calculation approaches, instead, requires the investigator to input a fixed number of days of treatment to be assigned either to the DUR(s) of interest or to each unit of the medicinal product package(s) of interest based, for instance, on prescribing or dispensing practices for the specific medicinal product and indication as applied in the country/region from which the study population was capture, or according to clinical or regulatory guidelines. Notably, days of treatment duration might be also already available in the data, e.g., as prescribed treatment duration or days supplied. Therefore, the fixed duration-based class was distinguished in two distinct calculation approaches:

1. fixed record duration, for which the investigator chooses to assign a fixed duration to the DUR(s) of interest (e.g., 30 days [28,29]). The DoT of each DUR of interest will be equal to the number of days of fixed duration inputted by the investigator, regardless of the medicinal product package characteristics, medicinal product package units or quantity of pharmaceutical product in the DUR;
2. fixed medicinal product package duration, for which the investigator chooses to assign a fixed number of DoT duration to each package of medicinal product in the DUR of interest (e.g., 30 or 60 days [28–30]), regardless of the number of units of presentation, the strength or the amount of pharmaceutical product.

### 3.2 The *CreateDoT* package

The R package *CreateDoT* was developed to run the *CreateDoT* function [25]. The primary output of the *CreateDoT* function was the number of DoT associated with DURs of interest calculated with the chosen RDD approach, while the secondary output was the average daily amount of active substance corresponding to the calculated DoT.

The input data consists of a *data.table* (R format) data set whose unit of observation is the DUR (see **Supplementary Table 2**). The function assumes that every DUR represents one prescription, dispensing or administration event corresponding to a defined amount of a specific pharmaceutical product or one or more unique medicinal product packages. If not already the case, prior to applying the function, the input data must be pre-processed so that there is one row per DUR.

Information available in a DUR may vary from one datasource to another. Consequently, information potentially useful as input data for the function, like PDD, medicinal product characteristics (e.g., unit of presentation, active substance, number of dosage units, strength, among others), or even prescribed treatment duration or days supplied, might be not always available or might be partially missing. It is the investigator’s responsibility to define which calculation approach must be applied and consequently which information available in the DURs of interest are to be used accordingly.

#### 3.2.1 Inputs

Input data can be distinguished in 1) parameters to be imputed based on specific assumptions of the investigator and 2) parameters to be retrieved from recorded data.

First, one of the five calculation approaches should be chosen and entered by the investigator. Therefore, in case a DD-based approach is chosen, the investigator should also input:

- a **Daily Dose (DD)**, defined as either an amount of active substance, an amount of pharmaceutical product or a defined number of units of presentation.
- the corresponding **DD unit** of measurement, which depends on the chosen DD. For example, the inputted daily amount of active substance might be measured in “mg” or “g”, the daily amount of pharmaceutical products might be measured in “g” or “ml”, while the daily number of units of presentation might be measured in “tablets” or “syringes”.

In case a fixed-duration approach is chosen, the investigator should input:

- **fixed days of duration**, i.e the number of days to be associated either with each DUR of interest or with each medicinal product package of interest contained in the DURs of interest.

#### 3.2.2 Outputs

The output of the function includes three new variables: *calculated_dot*, i.e., the calculated number of days of treatment associated with each DUR of interest; *calculated_dd*, i.e., the average daily amount of active substance calculated based on the corresponding DoT; and the *calculated_dd_unit*, i.e., unit of measurement of the *calculated_dd* (see **Supplementary Table 2**).

For each calculation approach, except the “amount of active substance per day”, where the originally imputed DD is already expressed as the amount of active substance, the function calculates the amount of active substance per day dividing the total amount of active substance contained in the DUR of interest by the calculated DoT. If the DUR of interest includes a medicinal product containing a combination of drugs (i.e., with more than one active substance, such as sacubitril/valsartan), the DD will be calculated for both active substances, whenever the necessary information is available.

In addition, two intermediate variables corresponding to the total active substance amount contained in DUR (*total_active_substance_amount_per_DUR)* and the corresponding unit of measurement (*total_active_substance_amount_per_medicinal_product_unit)* are also generated. They are necessary for both to be used by the function for the application of the DD-based approaches “active substance amount per day” and also to compute the calculated DD. The *total_active_substance_amount_per_DUR* is calculated by multiplying the active substance amount contained in one unit of presentation by the total number of unit of presentation contained in the DUR of interest (e.g., 50 * 50 mg = 2500 mg).

To calculate the total active substance amount per DUR, two different computational formulas can be used depending on the data available in the source data:

a. if the active substance amount per unit of presentation is available:

*total_active_substance_amount_per_DUR =* [*amount of active substance in one unit of presentation] x [number of units of presentation in the DUR]*

1. b. if the amount of the active substance contained in a medicinal product package is stored as a fraction or ratio of the pharmaceutical product:

*total_active_substance_amount_per_DUR = [fraction or ratio of the active substance of interest contained in the pharmaceutical product] x [amount of pharmaceutical product in one unit of presentation] x [number of units of presentation in the DUR]*

### 3.3 APPLICATION OF *CreateDoT*

#### 3.3.1 Preparatory steps

Before implementing the *CreateDoT* function, a few preparatory steps are necessary. A workflow describing the preparatory steps required before running the *CreateDoT* function are described in **Figure 1**:

1. the active substance(s) related to the exposures of interest should be defined (e.g., a list of ATC or RX norm codes or active substance names);
2. the selected active substance(s) should be mapped to the identifiers of the corresponding medicinal product packages or pharmaceutical products as stored in the reference dictionary. Such dictionaries are generally national (e.g., British National Formulary in the United Kingdom, the Dutch Z-index, among others) [31,32], although international initiatives are ongoing [33]. This step is optional as some data sources or exposure duration estimation strategies do not require this (e.g. in case a fixed-duration is assigned to each DUR containing the active substance of interest);
3. the resulting list of medicinal products identifiers should be linked to those recorded in the study data instance to retrieve the DURs of interest corresponding to the exposure drug identifiers’ codelist;
4. the variables of the DURs should be mapped to the universal concepts described in the RDD calculation approaches;
5. Finally, input parameters should be defined by the investigator, i.e. the calculation approach to be used with corresponding assumptions (e.g. DD value and unit, DUR duration or package duration) based on the study-specific scenario and corresponding context information (indication of use, recommended DD, recommended dosing schedule, dose strengths and package size available, expected treatment pattern, medicinal product package descriptors available in the dataset). Notably, the decisions of the investigator on the value of the input parameters can be taken considering different levels of granularity of DoT calculation (see **Table 3**):

1. active substance level, in this case the same assumptions and/or calculation approach are applied to any DUR containing the active substance of interest, (e.g., the WHO DDD of amlodipine corresponding to a DD of 5 mg [14] could be used to apply the calculation approach “active substance per day” to any DUR with oral amlodipine-containing medicine products; another example would be assuming a fixed duration of 15 days to any triptan-containing DUR);
2. at medicinal product package level, if assumptions and/or calculation approaches are defined according to the characteristics of each medicinal product package containing the active substance of interest, e.g., assume “DD=50 mg” to apply the calculation approach “active substance per day” to any medicinal product package of “TENORMIN® Atenolol 30 tablets of 50 mg” [26]; assume a “DD=5ml” to apply the calculation approach “amount of pharmaceutical product” to any medicinal product package of “TENORMIN® Atenolol 0.5 mg/ml injectable solution” [26]; assume “DD=1 tablet” to apply the calculation approach “unit of presentation per day” to any medicinal product package of “BLOKIUM-DIU® Atenolol/Chlortalidone 28 tablets of 100 mg/25 mg” [26];
3. at DUR level, if assumptions and/or calculation approaches are defined according to the information contained in each DUR of interest, e.g., since PDD is often available in prescription data, for each DUR with amlodipine-containing medicine the relevant PDD can be used to apply the calculation approach corresponding to the unit of measure of the relevant PDD, where DD=PDD.

**Figure 1.**
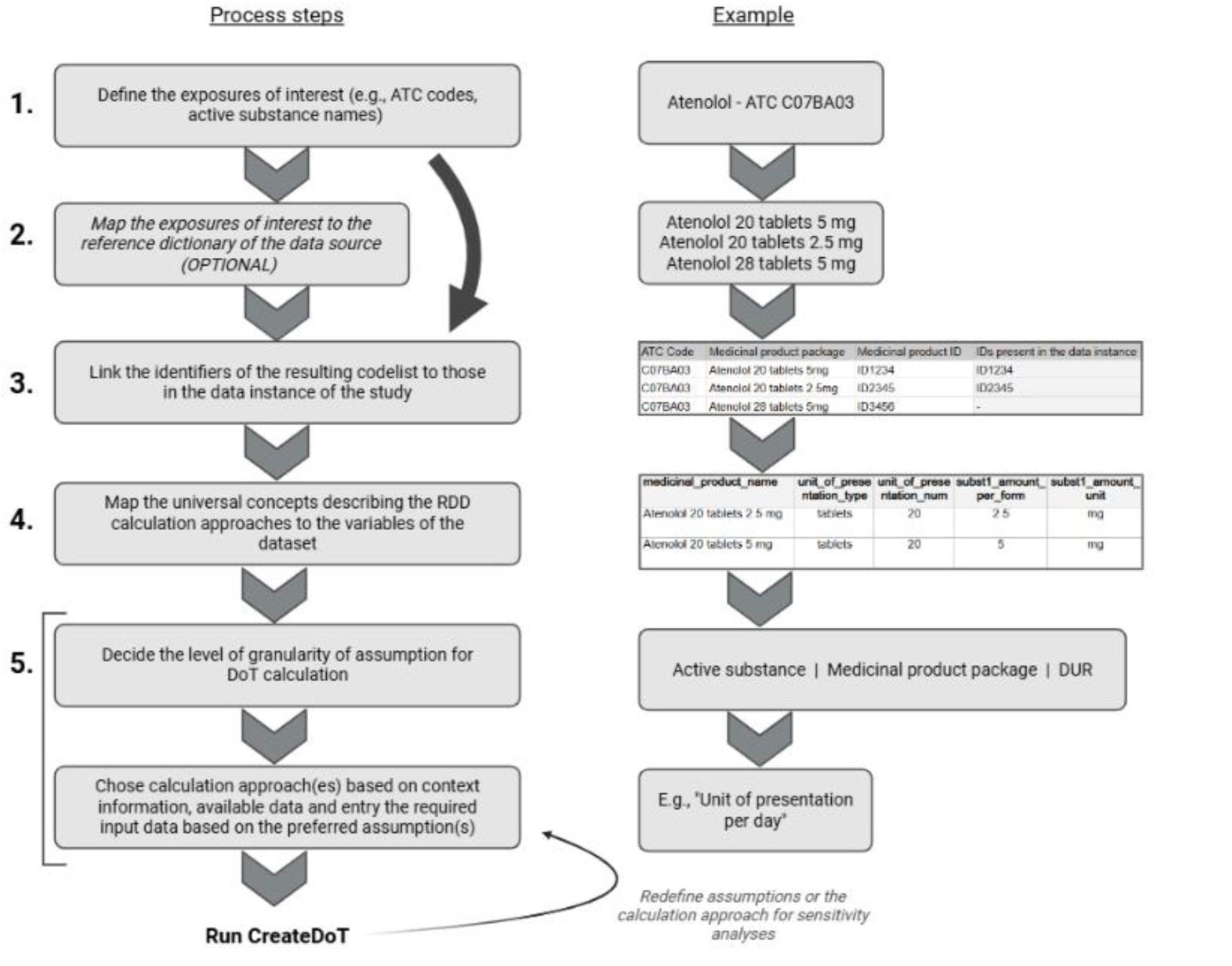
Workflow for the practical implementation of the CreateDoT function. *This figure has been created using* Biorender.com*. ATC: Anatomical Therapeutic Chemical; DoT: Days of Treatment; DUR: Drug Utilisation Record; RDD: researcher-defined duration*

**Table 3.** Level of granularity of assumption for DoT calculation.

| Level of granularity of assumption for DoT calculation | Description | Example |
| --- | --- | --- |
| Active substance | The same assumptions and calculation approach apply to all DURs containing the substance of interest | Apply "active substance per day" to any apixaban-containing DUR assuming DD=10 mg (based on DDD from WHO ATC classification) [14].<br>Apply "unit of presentation per day" to any simvastatin containing DUR assuming 1 daily tablet.<br>Apply "fixed record duration" to any acitretin containing DUR assuming 30 days duration. |
| Medicinal product package | Calculation approaches are tailored to each medicinal product characteristics | Apply "unit of presentation per day" to the medicinal product package of "sumatriptan 6 tablets of 50 mg" assuming 2 tablets per day. |
|  |  | Apply "unit of presentation per day" calculation approach to medicinal product "sumatriptan 6 mg, 2 prefilled syringes of 0.5ml" assuming 1 syringe per day. |
| DUR | Calculation approaches are tailored to information available in each DUR (i.e., medicinal product package characteristics, prescribed quantity of pharmaceutical product, prescribed daily dose, prescribed duration, etc) | Apply "unit of presentation per day", "active substance per day" or "pharmaceutical product per day" according to the unit of measurement of the corresponding PDD available in any DUR containing valproate. |

DD: daily dose; DDD: defined daily dose; DUR: drug utilisation record; PDD: prescribed daily dose; WHO: World Health Organisation

#### 3.3.2 Example case scenarios

With the purpose of illustrating the practical application of the *CreateDoT* function, we projected the universal concepts describing the RDD calculation approaches to the variables of the ConcePTION CDM [22,23] (see **Supplementary Table 3**), but any other data model can be used. Note that one DUR may contain multiple medicinal product packages.

In **Table 4** three case scenarios are reported as illustrative dummy examples of the application of different calculation approaches in the *CreateDoT* function. These examples are also available in the following publicly available repository in GitHub [34].

**Table 4.**
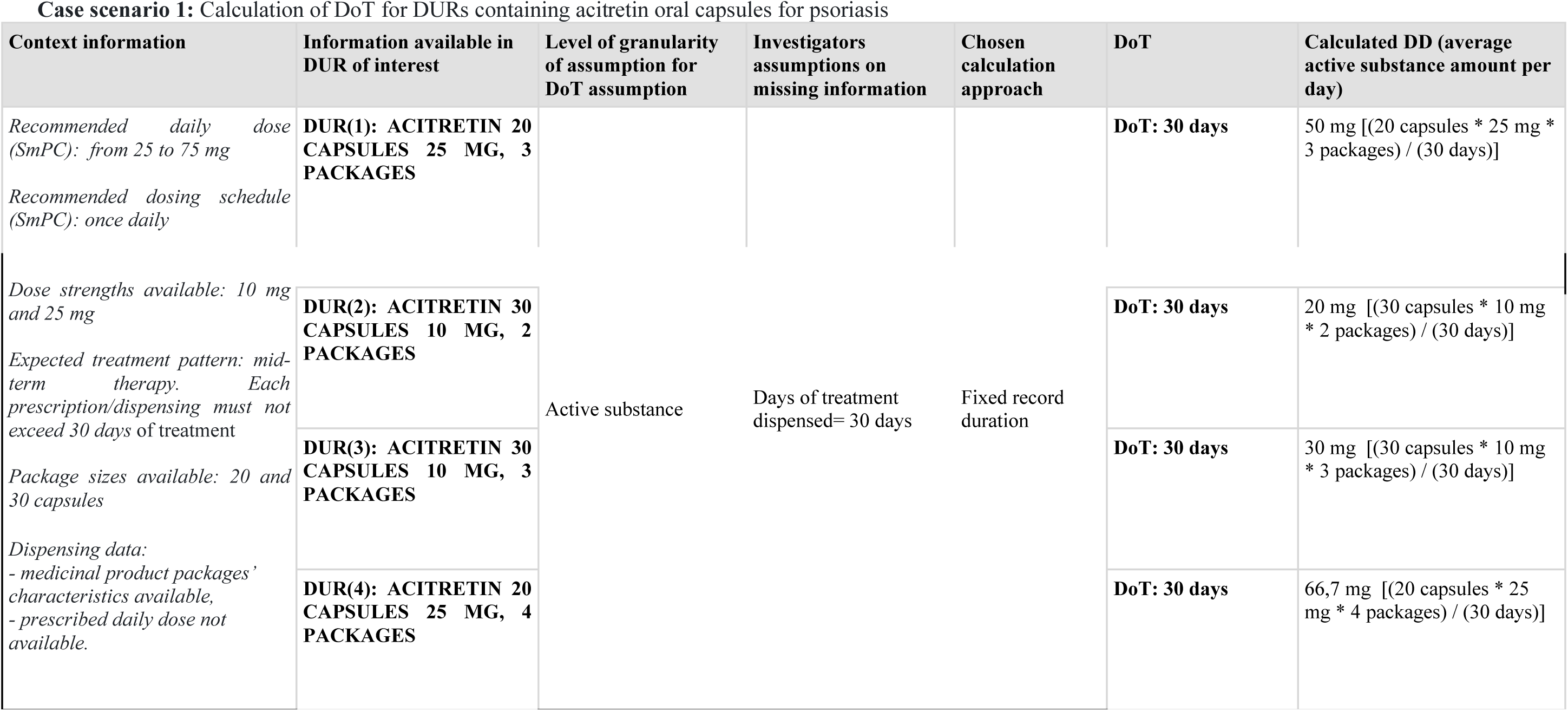
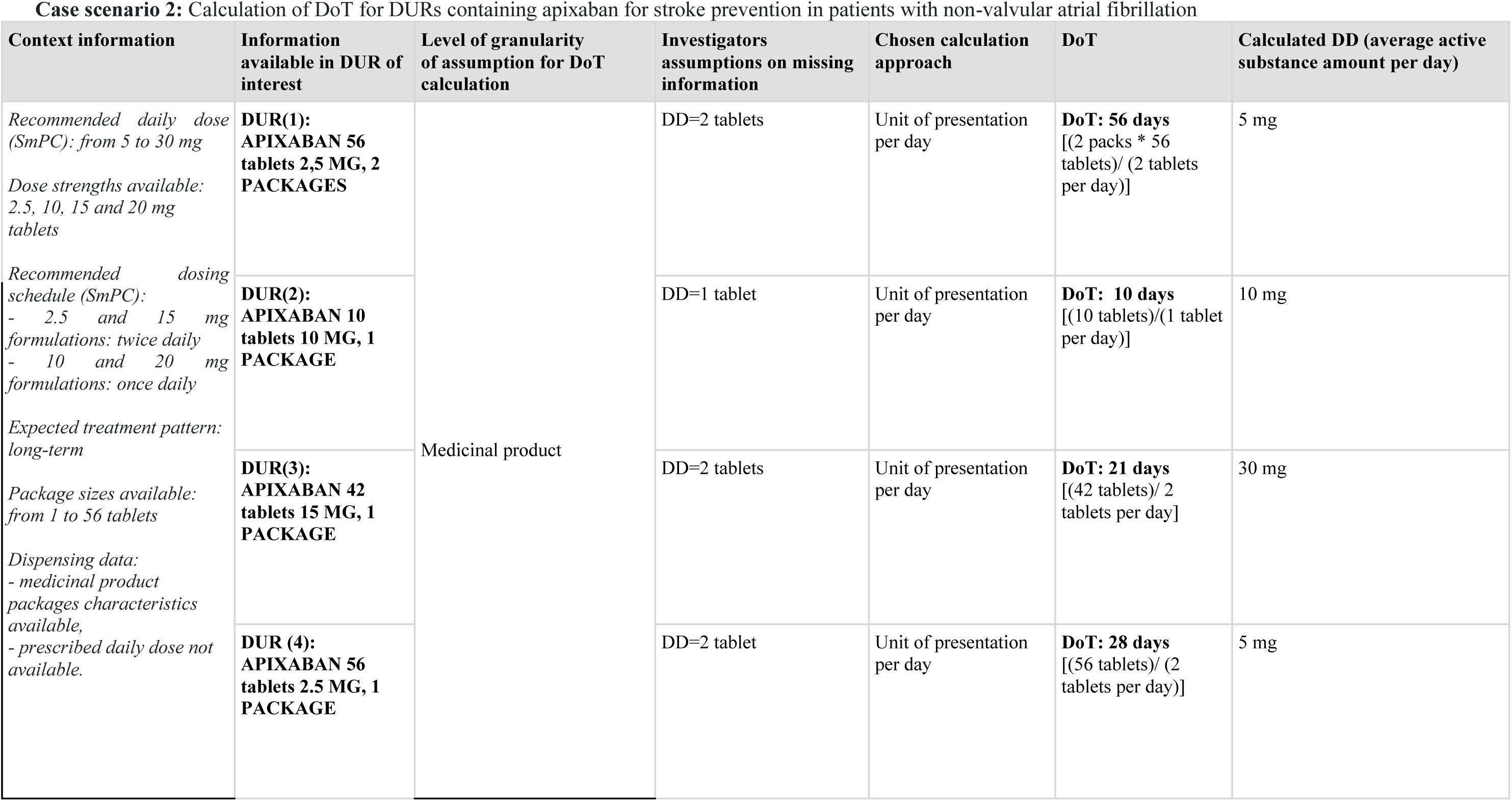
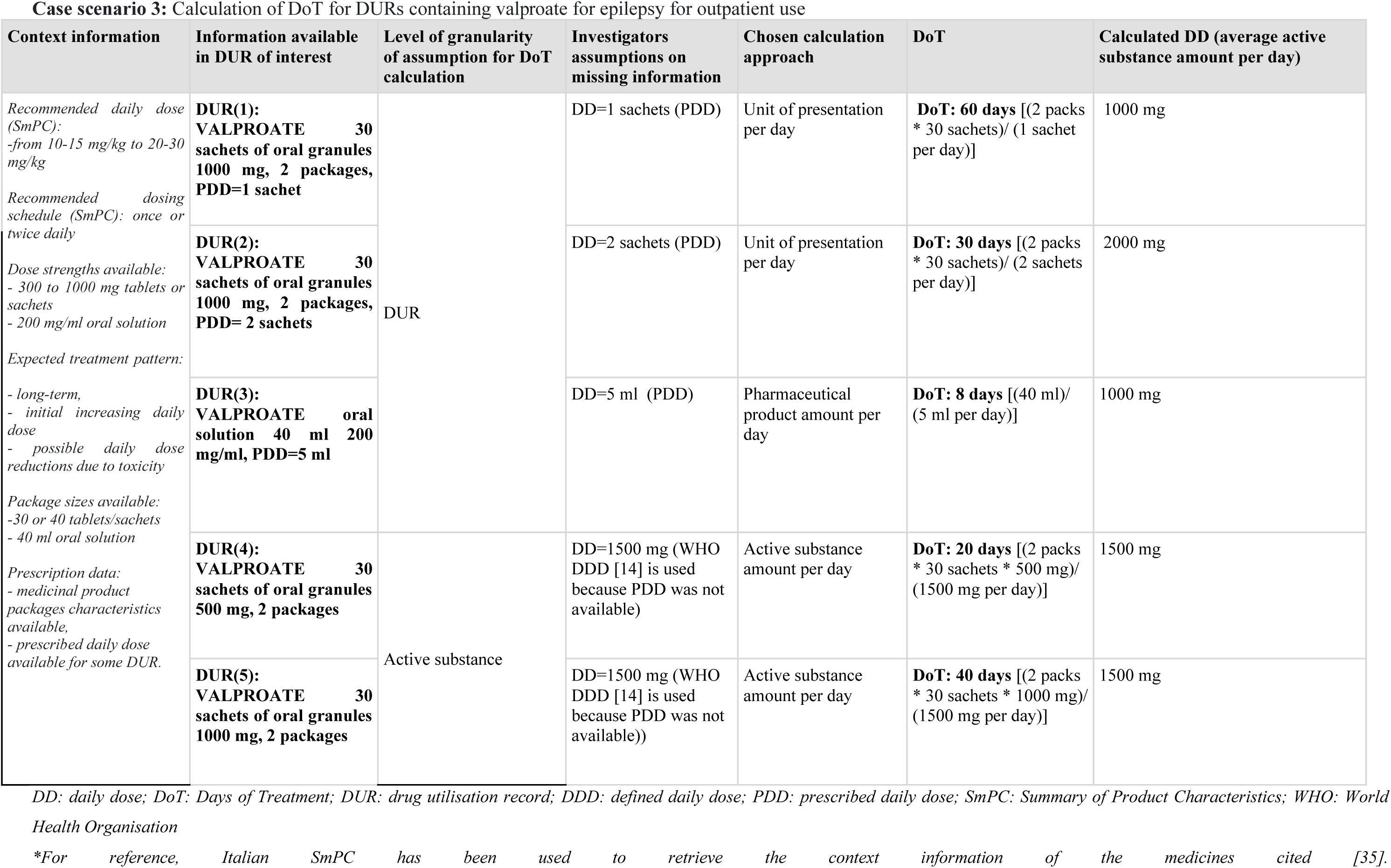
Examples of applications of Researcher Defined Duration-calculation approaches to different case scenarios [34]*.

## 4. DISCUSSION

In this study, RDD approaches for calculating days of treatment associated with electronic DURs were systematically classified. Based on such classification, the universal concepts useful as input for the relevant calculation formulas were identified and described using the EDQM and the ISO standard terminologies [20,21]. An open-source R function was developed to standardise the implementation of the different approaches for the calculation of DoT to be associated with single DURs with the final aim of facilitating documentation, transparency and reproducibility of methods in pharmacoepidemiological and drug utilisation studies. The input file structure itself represents a standard reporting template for documenting investigators’ assumptions and methodological choices adopted for DoT calculation.

### 4.1 Types of DURs and challenges for DoT estimation

DURs originated from prescribing or medication orders [36–39] often lack prescribed duration or discontinuation dates, and may not necessarily correspond to an actual dispensing or patient use of the drug of interest. Furthermore, discontinuation dates are rarely recorded, thus requiring the estimation of treatment duration based on prescribed quantities, dosing instructions, or refill intervals. DURs that capture dispensing events linked to reimbursement claims [36,37,40] provide more reliable information on the timing of exposure initiation compared to prescribing data and usually contain detailed qualitative and quantitative information on the supplied drugs. Although in some cases they also capture information on the supplied number of days of treatment, they usually lack clinical details, such as the intended dose regimen and indication, or whether the medication was actually consumed. DURs from disease or drug registries [36,41] may contain information on medicine administration (e.g., vaccination registries) or curated exposure information, sometimes verified by clinicians or through linkage with administrative dispensing records, but they often cover only specific patient populations or therapeutic areas and may lack information on comedications and data for extended periods of time, possibly impacting on the assessment of long-term exposures. Across all these DUR types, treatment start date is usually assumed to be the day in which the record is triggered (i.e., the prescription, dispensing or administration date) while the end date is usually not explicitly recorded and must therefore be assigned based on data available in the DUR and/or investigator’s assumptions on missing information [10,36].

Notably, the generalisation and systematic classification of RDD calculation approaches proposed in this study is intended to accommodate any RDD method that can be possibly used to calculate DoT using any type of DUR from real-world healthcare data sources. This allows for the standardisation of RDD calculation methods, which is crucial to ensure consistency of the analytical approach across studies and among data sources when multi-database studies are performed, and facilitates both the reporting of methods, the interpretation and comparability of study results. Several studies have compared the results obtained from the estimation of prescription/dispensing duration using either different sources for the assumed DD (e.g., medicine labels, clinical guidelines, the DDD from the WHO/ATC system, or fixed-durations with predefined time-windows) [7,9,12,42]. Although RDD calculation approaches demonstrate a high sensitivity [9], they do not offer a one-size-fits-all solution.

### 4.2 Strengths and limitations of the different RDD calculation approaches

Each RDD approach has distinct strengths and limitations which depend on the specific study scenario and the available information recorded in the study dataset. The investigator choices, with respect to the specific RDD approach to be adopted, the corresponding assumptions required and the level of granularity for its implementation, are crucial to minimise exposure misclassification. Since such choices are usually not validated and they might have a significant impact on study results [16–19], sensitivity analyses varying the chosen RDD calculation approaches and/or assumptions should be always considered and pre-specified in the study protocol to confirm the robustness of the primary methodological approach applied for DoT calculation.

DD-based calculation approaches require researchers to impute an assumed DD or use the PDD in case it is available in the dataset. However, their implementation requires both qualitative and quantitative information about the medicinal product (e.g., strength, type and number of units of presentation, active substance amount, concentration), which may not always be available. Moreover, conversion of units of measurement (e.g., aligning DDs and presentation forms such as tablets or mg) may also be necessary before implementation in order to harmonise units of measurement of the DD chosen by the investigator (e.g., 3 mg of paracetamol per day) with respect to descriptors of the medicinal product, as available in the DURs of interest (e.g., paracetamol 30 tablets, 1000 mg per tablet). Therefore, DD-based calculation approaches can be the preferred choice whenever PDD is available (more likely if our data source is an electronic health record database or a drug registry) [36–38,41], or if assumptions on DD are expected to be reliable (e.g., DD for statin is always 1 tablet per day), and the necessary descriptors to implement the calculation formulas are also available in the dataset (for example, in claims data sources timing and quantity of supplied medicinal product packages might be very detailed) [36,37,40].

In contrast, fixed duration-based calculation approaches offer a simpler alternative in terms of calculation formulas and information needed for their implementation compared to DD-based approaches. Fixed duration-based calculation approaches require the researcher to impute the assumed number of days of treatment to be associated with each DUR or medicinal product package of interest. Therefore, fixed duration can be preferred over DD-based whenever the assumption on the number of days of treatment to be associated with a DUR or a medicinal product package is considered more reliable than the assumption on the corresponding DD. For instance, fixed duration calculation approaches might be preferred when PDD is not available from the data source and, at the same time, the recommended DD for the medicines of interest might be highly variable between patients and/or unpredictable (e.g., triptans for migraine), and/or when prescription/dispensing duration is subject to local clinical guidelines, legislations or regulatory restrictions (e.g., retinoid or methylphenidate prescriptions/dispensings not exceeding 30 days [28,29], amlodipine not exceeding 60 days [30], dispensing of reimbursed chronic inexpensive drugs restricted to 3 months [43,44], etc). These calculation approaches might also be useful when the information about the dispensed number of medicinal product and/or medicinal product package information is unknown. Moreover, fixed duration approaches have the advantage of reducing the risk of calculation errors and can be applied even to datasets that lack qualitative and quantitative descriptors of medicinal products. However, these calculation approaches may oversimplify dosing regimens if not appropriately used.

### 4.3 Reporting of RDD-related methodological choices using the *CreateDoT* framework

The conceptual framework proposed in this paper emphasises the importance of researcher’s choices and assumptions with respect to DoT calculation, facilitating documentation and reporting of computational formulas, as well as of assumptions on DDs or fixed-time windows applied (see **Supplementary Table 2**). The present work also considers an additional dimension useful to accurately describe the methods applied for DoT calculation, that is the level of granularity at which methodological choices and assumptions are applied for DoT calculation (i.e., active substance level, the medicinal product level or the DUR level). It can be particularly relevant in case different step-wise RDD methods and assumptions are applied at different levels, depending on the information available (see below the discussion of the case scenario 3 reported in **Table 4**).

### 4.4 Examples of application of RDD calculation approaches

To better illustrate the application of the different RDD calculation approaches together with the proposed reporting framework, we described three dummy examples of DoT calculation. In the first example concerning the calculation of DoT for DURs containing acitretin oral capsules for psoriasis, a 30-days fixed-DUR duration assumption was applied at active substance level, i.e. to any DURs containing acitretin-based medicinal products. The decision was driven by the absence of PDD information, the wide inter-individual variability of the recommended daily dose, and the regulatory restriction of prescription/dispensing duration to a maximum of 30 days of treatment [26,28,35]. This is a clear example in which the chosen fixed-duration approach can be a priori considered as the most reliable approximation of the actual exposure in the study population.

In the second example, calculation of DoT for DURs containing apixaban for stroke prevention in patients with non-valvular atrial fibrillation, the DD-based approach “unit of presentation per day” was applied at medicinal product level, meaning that the assumption about the value of the DD (i.e., number of tablets) was defined according to the characteristics of each medicinal product of interest. This methodological choice was justified by the absence of PDD information (as it could occur in administrative/claims DURs) and the assumption that the DD could be reliably predicted based on tablet strength and posology recommendations [35].

The third example concerns the estimation DoT for DUR of oral valproates for epilepsy. In this scenario, the use of prescription data with PDD information available in only 3 out of 5 DURs was represented. In order to maximise the use of information available in the dataset, a step-wise application of RDD methods and assumptions was applied: whenever the PDD was available in the DUR, a DD-based approach was applied leveraging DUR level information on PDD; and where the PDD was not available, a DD corresponding to the DDD from the WHO ATC/DDD index (i.e., 1500 mg per day) [14] was assumed. In these cases, the “active substance per day” DD-based approach was applied at the active substance level to any DUR with a valproate-containing medicine and no PDD. It is important to note that DDDs [14] may not accurately reflect the actual PDD, although they are often used for DoT calculation [9,27,45]. Several studies have shown that DoT estimated using DDDs can significantly differ from those obtained using the PDD or other assumed DDs [7,9,12,42].

### 4.5 Applications of the CreateDoT function

In order to standardise the calculation methods corresponding to the application of the RDD approaches, we developed an R function [25] based on parameters corresponding to the universal concepts described using ISO and EDQM standard terminologies [20,21]. This approach resulted in the development of a data model-agnostic function. Notably, for the purposes of this paper we projected the function parameters to the ConcePTION CDM variables for illustrative purposes only, since the function is intended to be potentially applicable to any real-world data format. Detailed examples on the implementation of the CreateDoT function on dummy DUR in the ConcePTION CDM format, including R scripts, are available on GitHub [34]. The primary output of the *CreateDoT* function (i.e., *calculated_dot*) is represented by a number of DoT associated with one single DUR.

The function in itself does not compute a variable indicating the end of continuous treatment episodes consisting of more than one consecutive DUR. Therefore, in case episodes of continuous treatment have to be created, it is the responsibility of the user to make further assumptions on exposure start date, overlaps and gaps between the estimated duration of two or more consecutive DURs of interest [5,15,46]. Notably, the choices underlying such additional steps for defining episodes of continuous exposure are out of the scope of the present work.

The DoT calculated through the *CreateDoT* function can be used both as an analysis variable itself, or, for instance, as an input to build episodes of continuous treatment using other packages already available [46–49]. The primary output variable, in fact, may be used as input for *AdhereR* [47,48], which focuses on patient adherence and how well patients follow their medication schedules using metrics such as proportion of days covered or medication possession ratio. On the other hand, the output of the functions *doseminer* and *drugprepr* packages [15], which were developed to extract free-text drug prescription instructions in a structured form, may serve as input for the *CreateDoT* function.

### 4.6 Strengths and limitations

The main strength of this work is the generalisation and standardisation of RDD definitions. The proposed conceptual framework is intended to reduce heterogeneity and facilitate transparency of study methods reporting in pharmacoepidemiological studies and provide investigators with a simple and comprehensive classification of possible methodological options for DoT calculation which can be chosen according to the information available in a study dataset. Moreover, the use of EDQM [20] and ISO [21] standard terminologies to describe the universal concepts corresponding to the function parameters represents an additional strength. This approach potentially allows projecting and adapting the function to any type of dataset content and format. Another strength of the present work lies in the development of the function as an open-source R package, which enhances usability, promotes transparency, and facilitates feedback from the research community for continuous improvement.

The main limitation of this work is that practical implementation in a real study is still pending. However, application of the *CreateDoT* function is anticipated in upcoming pharmacoepidemiological and drug utilisation studies of the ConcePTION and EU PE&PV Network [50]. Another limitation of this study is that the identification of RDD calculation approaches and universal concepts was based primarily on expert opinion rather than a formal scoping or systematic review. Although, our experience suggests that it is unlikely that major alternative methods would have been missed. Finally, we did not address data-driven duration approaches such as PRE2DUP, Sessa empirical estimator, parametric gap modelling, machine learning methods or reverse waiting-time distributions [7,9,15,51,52], which represent important alternatives to the RDD approaches that deserve to be considered for the future development of the conceptual framework presented in this paper and the Create DoT function.

## 5. CONCLUSIONS

With this study we developed the *CreateDoT* framework intended to foster the harmonisation and standardisation of the classification, terminology, implementation and reporting of Researcher Defined Duration (RDD) calculation approaches used in drug utilisation and pharmacoepidemiological studies for calculating days of treatment associated with electronic Drug Utilisation Records (DURs). Conducting sensitivity analyses testing more than one choice is recommended to evaluate the robustness of results under different RDD assumptions. The application of the *CreateDoT* function will ensure consistency of the application of RDD calculation approaches across data sources, facilitating both documentation and reproducibility of study methods across different studies and among data sources participating in the same multi-database study.

## STATEMENTS AND DECLARATIONS

### Author Contributions

All authors contributed to the conceptualization and methodology of the study. All authors contributed to the creation of the formulas and interpreting their outputs and applicability. Judit Riera-Arnau, Olga Paoletti and Giuseppe Roberto were responsible for results curation and formal analysis. Rosa Gini and Olga Paoletti contributed to software development. Supervision and funding acquisition were undertaken by Rosa Gini and Giuseppe Roberto.The first draft of the manuscript was written by Judit Riera-Arnau and Giuseppe Roberto, and all authors commented on consecutive versions of the manuscript. All authors read and approved the final manuscript.

### Data Availability Statement

Data used for this manuscript, the CreateDoT function theoretical content and its related R package are stored in the following open access GitHub repository: https://github.com/IMI-ConcePTION/CreateDoT/wiki/.

### Declaration of generative AI

During the preparation of this work the author(s) used ChatGPT 4.1 as a supplementary tool to assist with content synthesis and language editing.

### Ethics Approval

Not applicable.

### Consent to participate

Informed consent was not required from the patients due to the nature of the study.

### Consent to publish

Not applicable.

---

**Supplementary Table 1.** Examples for the MEDICINES and PRODUCTS tables of the ConcePTION Common Data Model

**Supplementary Table 2.** Input and output template shell tables for CreateDoT implementation and reporting (illustrated using variables from the MEDICINES and PRODUCTS tables of the ConcePTION Common Data Model as example inputs)

**Supplementary Table 3.**
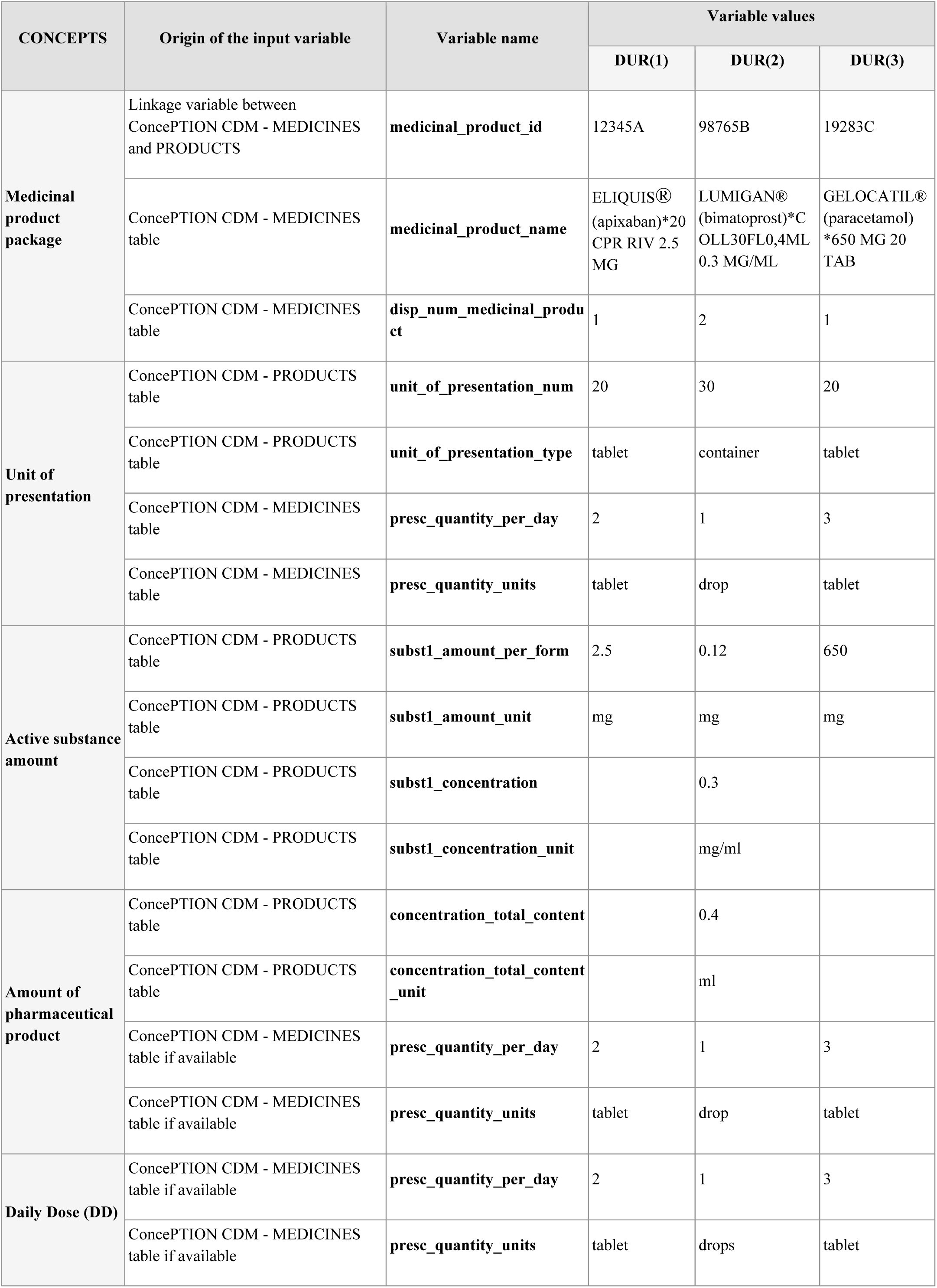
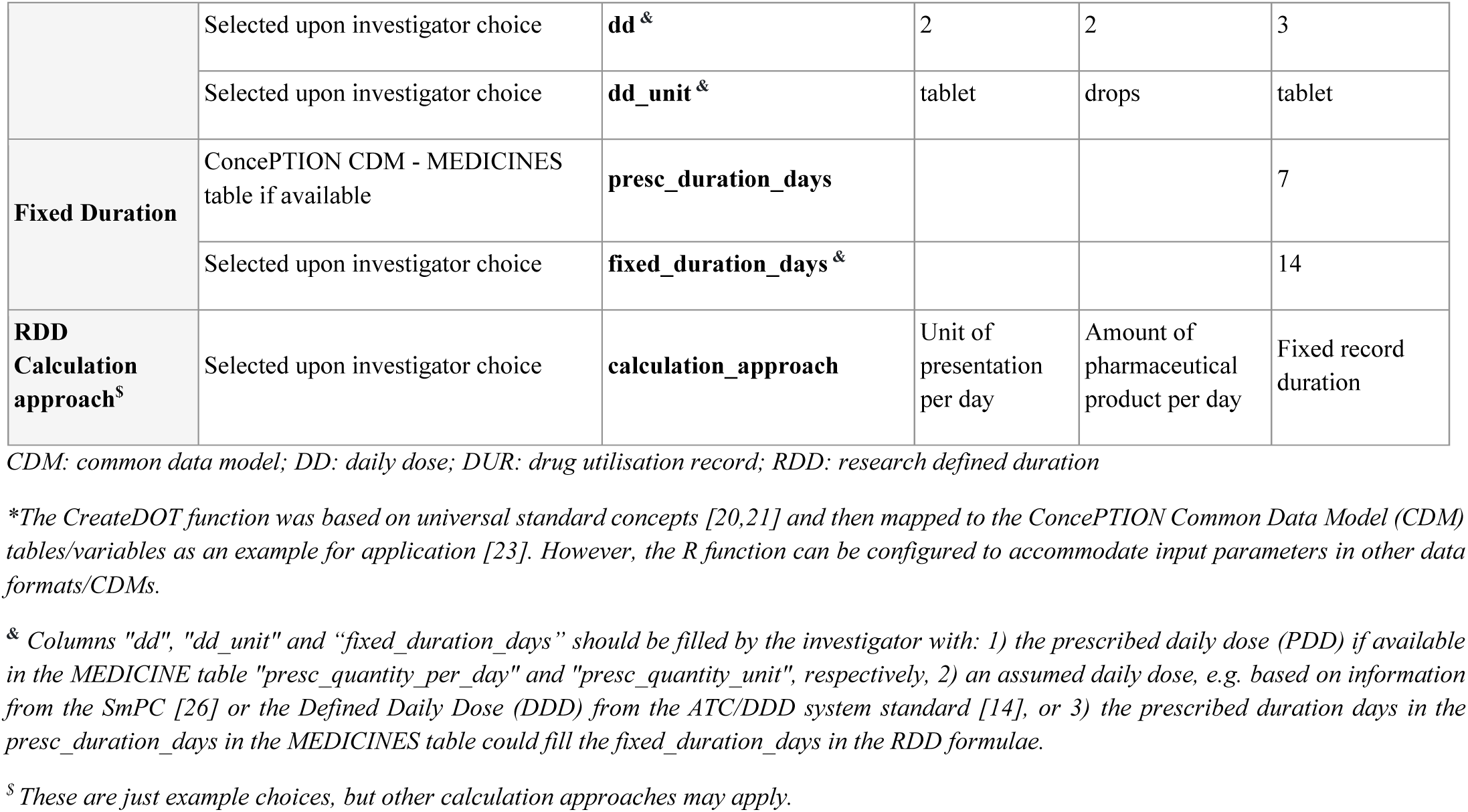
Projection of universal concepts to input parameters for the *CreateDoT* function application, using 3 dummy DUR in the ConcePTION Common Data Model variables as an example.

## LIST OF ABBREVIATIONS

ATC: Anatomic-Therapeutic-Chemical code
CDM: Common Data Model
DoT: Days of Treatment
EDQM: European Directorate for the Quality of Medicines
DD: Daily Dose
DDD: Defined Daily Dose
DUR: Drug Utilisation Records
IDMP: ISO standards for the Identification of Medicinal Products
ISO: International Organisation for Standardisation
PDD: Prescribed Daily Dose
RDD: Researcher Defined Duration
WHO: World Health Organisation

